# Time trends in cardiac doses in 10,000 patients receiving curative thoracic radiation therapy between 2009 and 2020

**DOI:** 10.1101/2024.02.18.24303007

**Authors:** Nora Forbes, Cynthia Terrones-Campos, Abraham Smith, Joanne Reekie, Sune Darkner, Maja Maraldo, Mette Pøhl, Signe Risumlund, Lena Specht, Soren M. Bentzen, Jens Petersen, Ivan R. Vogelius

## Abstract

**Background and purpose:** In the last 20 years, it has become well-documented that incidental cardiac exposure to ionizing radiation is associated with a clinically relevant increased risk of cardiovascular morbidity. In parallel, radiation therapy technologies have been developed to provide target dose coverage with less exposure to adjacent organs at risk. In the current work, we investigate trends in cardiac exposure among patients treated with curative intent radiotherapy from a single institution between 2009 to 2020.

**Materials and methods:** 10,215 treatment courses were analyzed from 9,966 patients treated with curative intent for intrathoracic or breast cancers in the period 2009-2020. All hearts were re-delineated using an AI model to ensure consistency over time. Cardiac doses were extracted in 3D from the record-and-verify system and converted, voxel-by-voxel, to equi-effective doses in 2 Gy fractions (EQD2) using α/β=2 Gy. Mean heart dose (in EQD2) and volume exposed to 5 and 40 Gy (V5 and V40Gy), respectively, were extracted. Time trends in these cardiac dose-volume metrics were investigated for each diagnosis.

**Results:** Patients with esophageal cancer had the highest mean heart dose (median = 11.67 Gy; IQR = 2.85, 18.18), while the lowest was observed in patients with breast cancer (median = 0.60 Gy; IQR = 0.30, 1.08) and lymphoma (median = 0.01 Gy; IQR = 0.00, 0.38). A decreasing trend over time was seen most clearly for patients with esophageal and lung cancers (p<0.05). Among patients with breast cancer, V40 decreased from 2009-2015 after which we observed an increase.

**Conclusion:** There has been a significant reduction in radiation exposure to the heart in patients treated in the period 2009-2020, likely due to increased awareness of cardiovascular toxicity and technological developments. The study also found a significant increase in V40 from 2015 to 2020 for patients with breast cancers, possibly related to increased prioritization of target coverage.

**Highlights:**

- 10,215 radiation therapy plans extracted and analyzed
- Hearts were re-delineated with machine learning on the therapy CT-scan to ensure time-independent delineations
- Cardiac doses were found to decrease over time, especially high dose exposure of intrathoracic lesions
- An unexpected increase in cardiac high dose volume (V40Gy) for breast cancer treatment plans in recent years was observed

## Introduction

The heart used to be considered a relatively radioresistant muscular structure, but publications in the modern era have demonstrated a clear association between cardiac exposure and risk of subsequent cardiovascular morbidity (Cutter et al., 2015; Darby et al., 2005, 2013; Van Nimwegen et al., 2016; van Velzen et al., 2022).

As a consequence, a number of technical developments have been introduced with the aim of minimizing the exposure of the heart to radiation. Examples of techniques with a documented dosimetric benefit during conventional exposure include proton therapy (Hassan et al., 2023) and deep inspiration breath hold (Nissen & Appelt, 2013; Pedersen et al., 2004; Petersen et al., 2015), whereas Intensity modulated radiotherapy (IMRT) has been shown to decrease cardiac exposure, but possibly at the expense of an increase in low dose bath (Chun et al., 2017).

Even for patients with a high risk of cancer recurrence, there have been reports of survival detriments with higher cardiac exposures (Atkins et al., 2021; Brink et al., 2022). Conversely, the Danish Breast Cancer Collaborative group showed that omitting an elective target to decrease cardiac dose may be unacceptable in terms of treatment efficacy (Thorsen et al., 2016a).

It remains to be defined to what extent increased awareness and technological advances allowing for more conformal treatment have impacted cardiac exposure in clinical practice. This study investigates trends in the cardiac dose received by patients treated with curative intent radiotherapy at a tertiary hospital in Denmark over the last decade (2009 to 2020). This study utilizes a previously presented method and pipeline for analyzing large cohorts of patients with detailed individual 3D dosimetry data and consistently delineate hearts (Smith et al., 2022). The study aim is to elucidate changes in dose delivered to the heart over time, to display the cumulative impact of gradual improvements in radiotherapy techniques and concerted efforts to reduce heart irradiation.

## Methods

We included all patients receiving curative intent radiotherapy at Copenhagen University Hospital - Rigshospitalet, Denmark between 2009 and 2020 with breast cancer, esophageal cancer, lymphoma, non-small cell lung cancer (NSCLC), or small cell lung cancer (SCLC), cf. Figure 1. Patients were identified in the ARIA record and verify system by local radiation therapy codes for treatments with curative intent. We extracted CT scans and 3D dose matrices using the Digital Imaging and Communications in Medicine (DICOM) protocol for automated access. Dose matrices obtained from the system represented dose calculations based on Anisotropic Analytical Algorithm or Acuros as employed at the time of treatment. Finnegan et al. showed that the guidelines and practice of contouring the heart have changed between 2009 and 2014, and this in turn will affect the dose-volume metrics for heart (Finnegan et al., 2020). Therefore, we used an in-house and open-source AI software, RootPainter3D, which facilitates the training of a 3D U-net model via corrective-annotation (Smith et al., 2022), to redelineate all hearts. Our cardiac delineations model was trained on 933 manually corrected CT scans from a prior immune toxicity study (Terrones-Campos et al., 2023). This model was then used to delineate all 14,044 CT scans within the entire population, yielding cardiac delineation on each CT scan. For quality assurance, all delineations were reviewed for gross errors in 2D with an in-house review tool (Figure A1). The average time to review each delineation was less than 5 seconds, which equated to approximately 16 hours of work. Questionable cases were extracted in 3D and assessed with a physician to determine inclusion.

**Figure 1.**
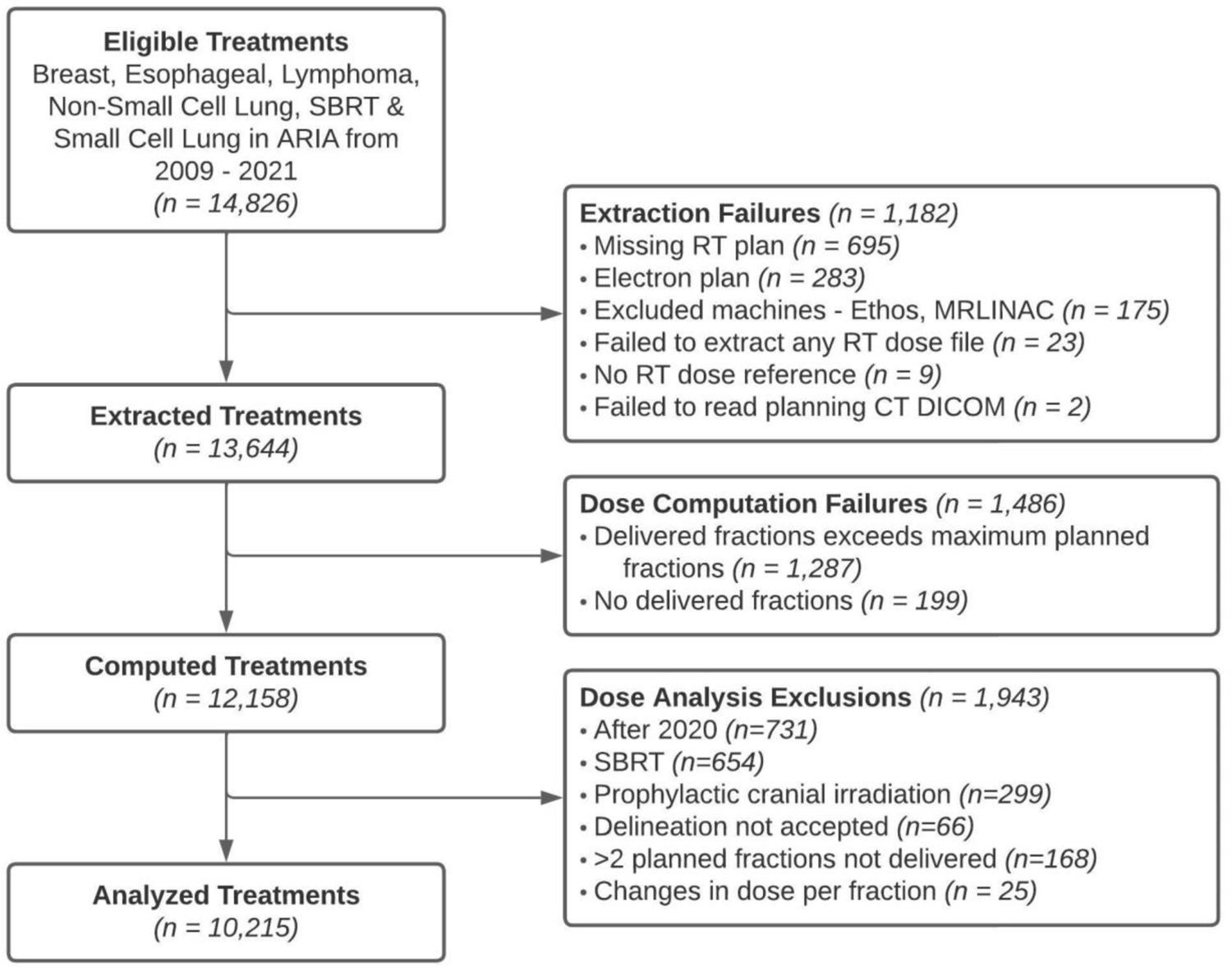
Consort diagram.

Fraction delivery times and dates were extracted from the record and verify system to aid dose assessment. When only one plan existed, the total dose was calculated as the sum over the fractions delivered. If two or more plans existed based on the same CT scan, the total dose was calculated as the sum over the total delivered fractions for each plan and then added together. If multiple plans existed based on different CT scans, it was not possible to directly sum fractions. In such cases, the plan with the most delivered fractions was used and we scaled up the dose to correspond to the number of fractions delivered in total across all CT scan/treatment plans. Cases with multiple plans, changes in treatment schedule, or otherwise not possible to reliably process as above were excluded. Similarly, we excluded electron plans and a small number of cases treated on an MR accelerator or Ethos as these systems could not be accessed automatically.

Fractionation correction was performed using Withers formula for equi-effective dose in 2 Gy fractions (EQD2) voxel-wise on the 3D dose matrix:

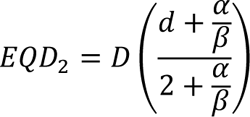

where D was the total dose at that voxel, d was the fraction dose at that voxel (D/fractions), and α/β was set to 2 Gy.

The mean heart dose (MHD) was calculated in EQD2. The dose volume histogram (DVH) values for each 1-Gy bin were extracted using EQD2 and calculated in cubic centimeters. We focused on one low dose-volume metric (V5 – volume receiving at least 5 Gy) and one high dose-volume metric (V40 - volume receiving at least 40 Gy).

Outliers in dose (several metrics) for all diagnoses were reviewed manually and compared to the record and verify system doses together with a medical physicist and/or oncologist to ensure that the automated dose extraction was accurate. This led to some necessary exclusion criteria to avoid erroneous cases (for example changes from curative to palliative intent).

The density of MHD was plotted for each of the patient groups. Scatter plots and trend plots were generated for each metric. Spearman’s Rho correlation was calculated for each metric and for each group.

R (R Core Team, 2023) was used within the RStudio environment (Posit team, 2023) to conduct analysis and generate figures along with the ggplot2 (Wickham, 2016), tidyverse (Wickham et al., 2019), and gridextra (Auguie, 2015) packages.

## Results

14,826 treatment courses (denoted “treatments” from here) on 13,855 patients in ARIA were eligible for this study, as shown in the consort flow diagram (Figure 1). Of these, 10,215 treatments on 9,966 patients could be included in the final analysis. Details of reasons for exclusion are given in the appendix, but in short, a combination of multiple scans/plans, “empty” courses and miscodings were common reasons.

10,215 treatments were included in the final analysis (Figure 1). Of these, the majority of treatments were patients with breast cancer (70%) and the smallest group was patients with SCLC (2%) (Table 1). The most common fractionation schemes are reported in Table 2.

**Table 1.**
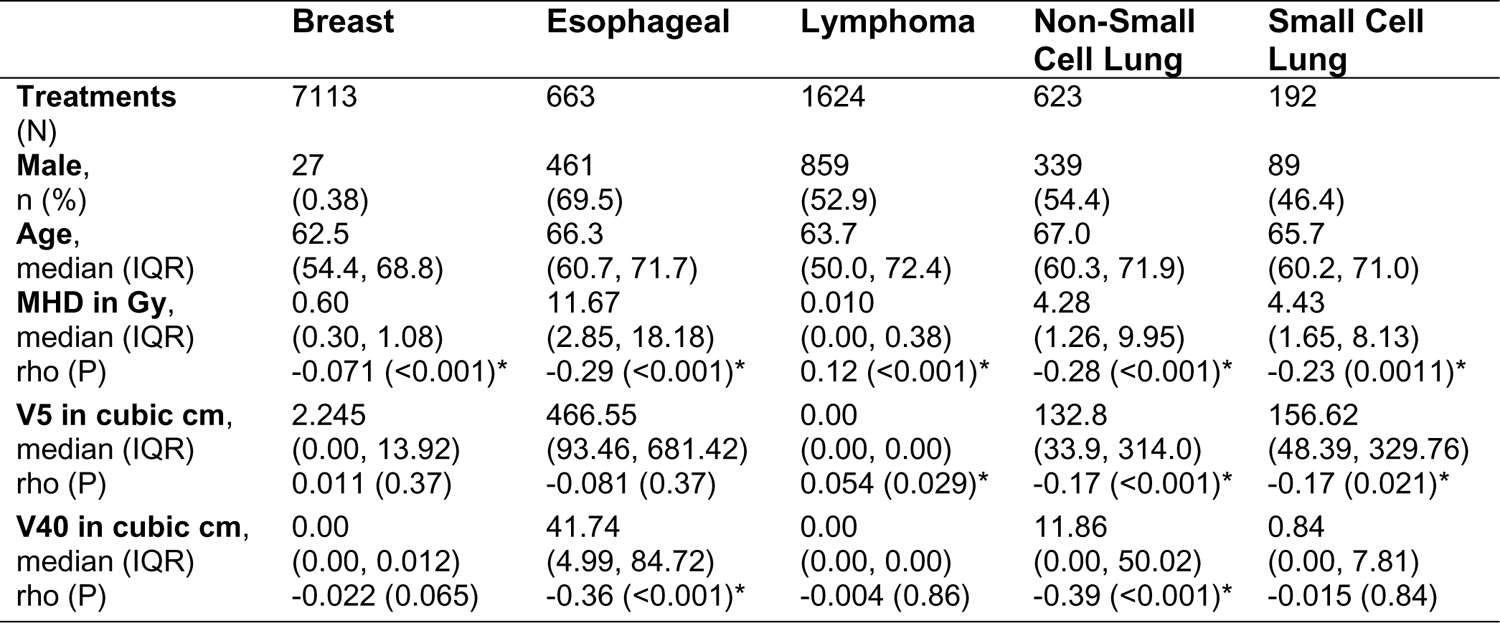
Demographics and dose statistics, including mean heart dose (MHD), heart volume receiving at least 5 Gy (V5), and heart volume receiving at least 40 Gy (V40).

**Table 2.**
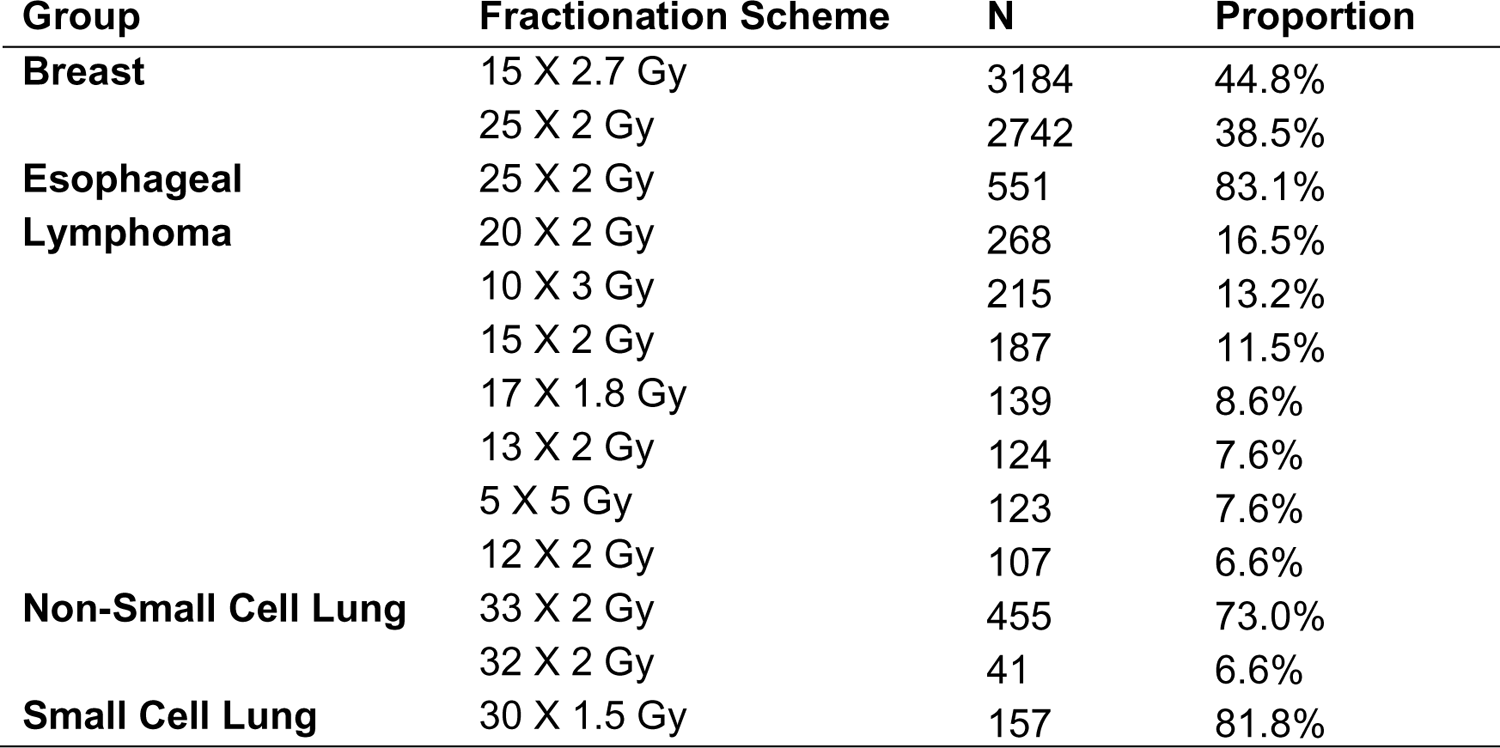
Fractionation schemes with at least 5% of each analysis group.

We observed a decrease over time in the annual number of patients receiving radiotherapy at Rigshospitalet for the indications considered here. This change reflects the transition of a satellite center to become a separate clinic causing a change in uptake area.

The dosimetric results are shown in Figures 2-4 with further details in supplementary figures. Starting with breast cancer, 92% of patients had a MHD below 2 Gy (Figure 2). The median MHD, V5, and V40 was second lowest across all analysis groups, while the means were the lowest (Table 1). The median MHD decreased significantly across the timeframe (p < 0.001) (Figure 3, Table 1). Neither the median V5 nor V40 decreased significantly (p = 0.37 and p = 0.065, respectively) (Figure 4, Table 1). High dose outlier cases decreased for both V5 and V40 (Figure A2). However, in the second half of the time period, the median MHD, V5, and V40 of breast cancer patients increased significantly (p < 0.001).

**Figure 2.**
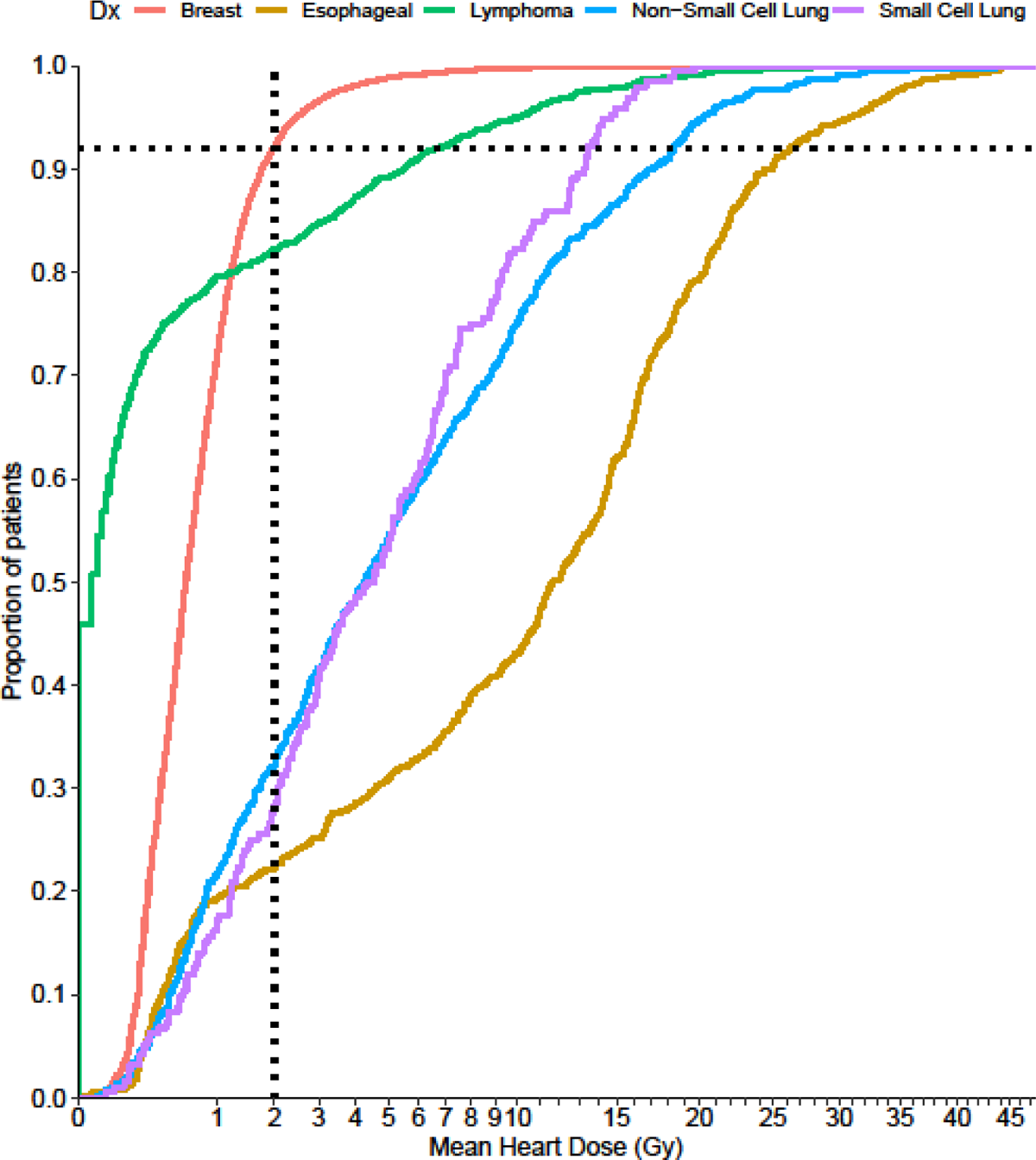
Cumulative density plot showing the proportion of patients by mean heart dose (MHD) in each analysis group. For example, approximately 92% of treatments in the breast cancer group received less than 2 Gy in MHD. The X-axis is scaled by a square root transform for clarity across the range of doses. Dose is reported in equivalent dose in 2 Gy fractions.

**Figure 3.**
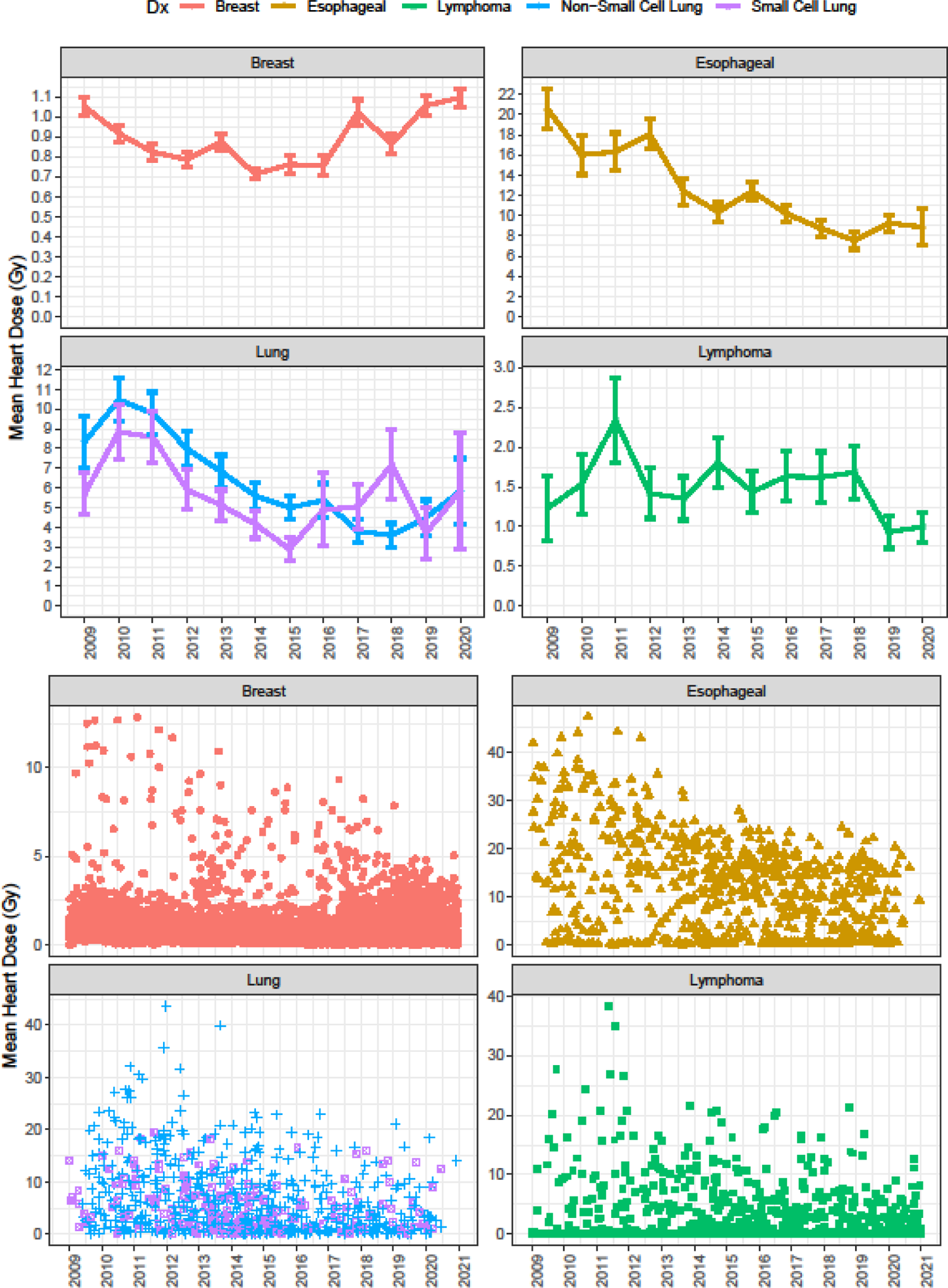
Annual trend plots (above) of median mean heart dose (MHD) for each analysis group, with lower (Q1) and upper (Q3) bounds. Scatter plots (below) of MHD for each analysis group. Note that each panel has an independent y-axis scale, and in particular the breast cancer scale is relatively small.

**Figure 4.**
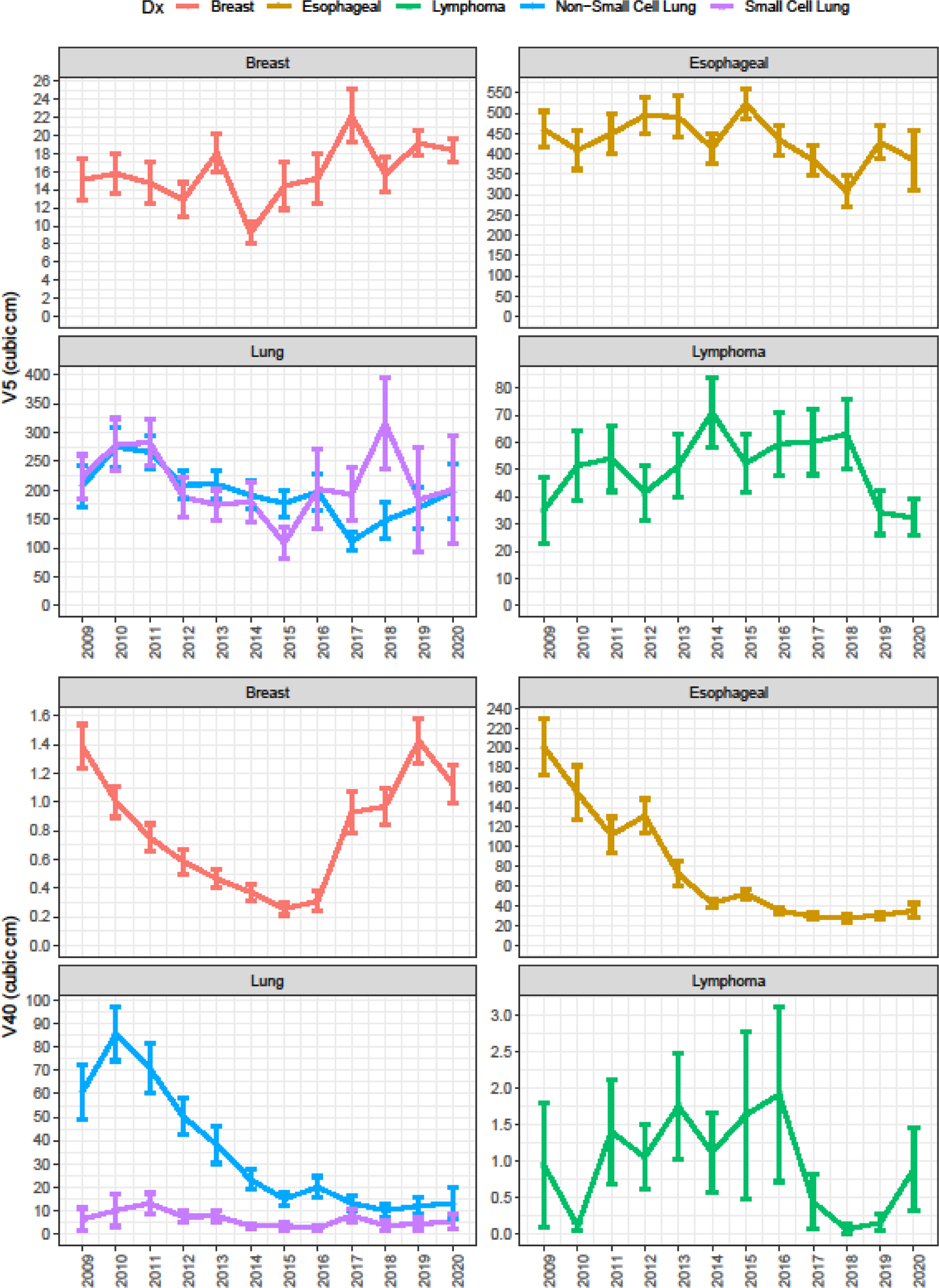
Annual trend plots of median heart volume receiving at least 5 Gy (V5) and heart volume receiving at least 40 Gy (V40) for each analysis group, with lower (Q1) and upper (Q3) bounds. Note that each panel has an independent y-axis scale, and in particular the breast cancer scale is relatively small.

Patients with lymphoma did not have a significant decrease in heart exposure over time (Table 2, Figure 3). Rather, there as a weak trend toward an increasing dose, which was driven by a decrease in cases with no heart exposure. Specifically, in the first 3 years 57% of MHD were zero, while in the last 3 years this fell to 38%. Mean MHD was 1.5 Gy, with 82% of the patients exposed to less than 2 Gy and 95% of the patients exposed <10 Gy.

Patients with Esophageal cancer had the highest MHD, V5, and V40. Only 22% of esophageal patients received less than 2Gy MHD. This is where we see the largest improvement in cardiac doses over time, especially V40 is reduced substantially. Specifically, median MHD decreased from 18.1 Gy in the first 4 years to 12.0 Gy in the last for years for the esophageal group. For NSCLC, median MHD decreased from 7.0 Gy in the first 4 years to 3.5 Gy in the last 4 years. The corresponding decrease for SCLC was from 6.9 Gy to 3.0 Gy. The dose reduction in MHD is driven by the decrease in the high dose exposure. We also observe that the occurrence of high dose outliers decreasing over time (Figure 3, Figure A2). Only patients with SCLC and NSCLC had a significant decrease in V5. Only patients with Esophageal and NSCLC had a significant decrease in V40.

## Discussion

Using an AI method, we were able to assess the MHD, and cardiac V5 and V40 of 10,000 patients treated with curative intent radiotherapy from 2009 to 2020 in key diagnostic groups. This real-world data analysis provides the end-result of cardiac exposures because of all combined changes of treatment in the study period, with a time-independent cardiac delineation method.

Starting with breast cancer the key findings were that cardiac exposures among patients with breast cancer in the Danish cohort were much lower compared to historical data, including the patients studied by Darby et al. (Darby et al., 2013). According to Darby et al., a MHD increase of 2 Gy is associated with a 14.8% excess relative risk of major coronary events. For a patient exposed to 2 Gy at age 50, this excess relative risk converts to an excess absolute risk of death from ischemic heart disease of just 0.1 % at attained age of 70 years (Calculated from cumulative incidence data derived from Darby et al supplemental table 13). Given that 92% of patients have lower exposure than 2 Gy, our data supports that cardiac risk pertaining to modern radiotherapy for most breast cancer patients is extremely low.

The Danish Breast Cancer Collaborative Group, DBCG, published the results of a population-based cohort study on the effect of internal mammary node irradiation in early node-positive breast cancer in 2016 with the main conclusion that omission of irradiation to spare the heart may negatively affect survival and disease control (Thorsen et al., 2016b). The DBCG data was also included in a recent seminal work of the early breast cancer trialists’ collaborative group (EBCTCG) individual patient data meta-analysis of 14,324 participants in clinical trials. The EBCTCG data shows an excess non-cancer related mortality in older trials when adding regional node irradiation, but this effect was absent in trials accruing from 1989-2013. In the more recent trials, a benefit was seen in reduction of breast cancer mortality without an adverse effect on non-breast cancer mortality. More specifically, the EBCTCG analysis report an absolute risk reduction in breast cancer mortality of 3% (95% CI: 1.1-4.9%) at 15 years and a risk reduction of 3% (95% CI: 1.0-5.0 %) for overall mortality when adding regional node radiotherapy. These numbers compared to the estimated 0.1% excess absolute risk of cardiac mortality clearly support for the increased prioritization of coverage of the target for breast cancer patients introduced in national and departmental guidelines after the IMN study in 2016. This clinical prioritization may explain the slight increase in cardiac exposure observed in recent years.

Nevertheless, continued monitoring, as presented here, can still point to continued optimization potential. Future work includes omission of radiotherapy for select patients (e.g., NCT03646955) or advanced techniques, such as proton therapy (e.g., ISRCTN14220944). It should be noted that the continued potential to gradually reduce high doses in outlier cases in recent years may imply that patient volume estimates for proton therapy trials using historical data, as in the DBCG proton trial, may overestimate the frequency of proton therapy indications for future patients (Stick et al., 2021).

The lymphoma patients are a quite different cohort, where the disease localization is highly heterogeneous. Lymphoma cases with high dose likely have a mediastinal target at the level of the heart, and this will effectively drive the incidental dose to the heart. DIBH was available as a treatment option from the early years for this treatment group (Rechner et al., 2017) which reduces cardiac exposure, but it appears that further improvement is difficult with current available methods. However, increased conformity to target should still be sought, including adaptation of the plan to daily patient anatomy and proton therapy as possible avenues for dose reductions to the heart.

We now turn to discuss lung and esophageal cancers. Among the intrathoracic solid cancers cardiac exposures vary considerably depending on target position. The observed reduction in MHD is driven by the reduction in high dose regions which is consistent with the increased high dose conformity and increased low dose bath associated with intensity modulated techniques (Bergom et al., 2021).

Additionally, the reduction is accompanied by reduced number and severity of high dose outlier cases. IMRT and volumetric arc techniques are used routinely for these patients in the later years. Deep inspiration breath-hold techniques have been evaluated for these patient groups but are not yet uniformly offered and do depend on patient compliance (Josipovic et al., 2019). Thus, there appears to be potential for further improvement in these patient groups. Central versus non-central location and mediastinal lymph node involvement will be important factors for individual exposure. Knowledge based planning is one way to individualize the expected achievable dose plan and the present work could be used as an avenue to increase the training data from the current standard of ∼100 patients (Li et al., 2017).

The decrease in patient volume at Rigshospitalet is due to a change in treatment center capacity in our historical uptake area. While there may be some differences in the patient population (e.g., socioeconomic status and lifestyle) it is not expected that this would have an association with the severity of cancer nor prescription of dose. Therefore, we do not anticipate this change in demographics to yield relevant bias to our conclusions.

Machine learning models for structure delineation combined with modern archival systems means that large-scale dosimetric analyses have been made possible as also demonstrated previously in breast cancer trial datasets (Finnegan et al., 2020). In the current study we used such methods to documents trends in incidental cardiac irradiation in a variety of diagnoses and over a period of 12 years in a single institution and in a very large patient cohort. Unfortunately, it is not possible to draw a specific causal relationship between individual changes in guidelines and the decrease in cardiac dose, however this serves as an overarching description of the combined impact of all these changes. Data points to improved cardiac sparing over time. A formal health economics perspective is outside of the scope of this paper, but we still find it relevant to emphasize that the employed techniques in later years (IMRT, dose planning expertise, awareness, and breath hold) come at a very modest – if any – excess resource expenditure per patient once the investment in equipment has been made.

The scalability of ML algorithms, of which the present study is one example of several, is a potential game changer in several areas (Vogelius et al., 2020). Here we demonstrated the ability to closely monitor doses to organs at risk across disease sites and over time in an unselected patient cohort. We were able to monitor detailed dose-volume data, including low dose bath and high dose exposure to the heart. The methods could be combined with machine learning techniques to account for the 3D heart exposure, one could generate much larger “knowledge-based planning” cohorts or the data could be coupled to registry or claims-based outcome data to elucidate new knowledge of rare, high-grade toxicities.

In conclusion, the analysis of a large consecutive dataset of patients documented continuous improvements in cardiac sparing over time consistent with continued technical improvements as well as clinical awareness.

## Data Availability

Data produced in the present study are available upon reasonable request to the authors

## Acknowledgments

We report institutional research and teaching contracts with Brainlab, Varian, and ViewRay. This project has received funding from the European Union’s Horizon 2020 research and innovation programme under the Marie Skłodowska-Curie grant agreement No 801199. The authors acknowledge support from Kirsten and Freddy Johansen’s award and National Cancer Institute (Grant No. P30 CA 134274-04). SMB was supported by funds through the Maryland Department of Health’s Cigarette Restitution Fund Program. IRV and JP received research funding from Varian Medical Systems. IRV and JP are supported by Danish Cancer Society (Grant No. R231-A13976). This work is supported by Danish National Research Foundation (DNRF) grant number 126 (PERSIMUNE).

## Appendix

**Figure A1.**
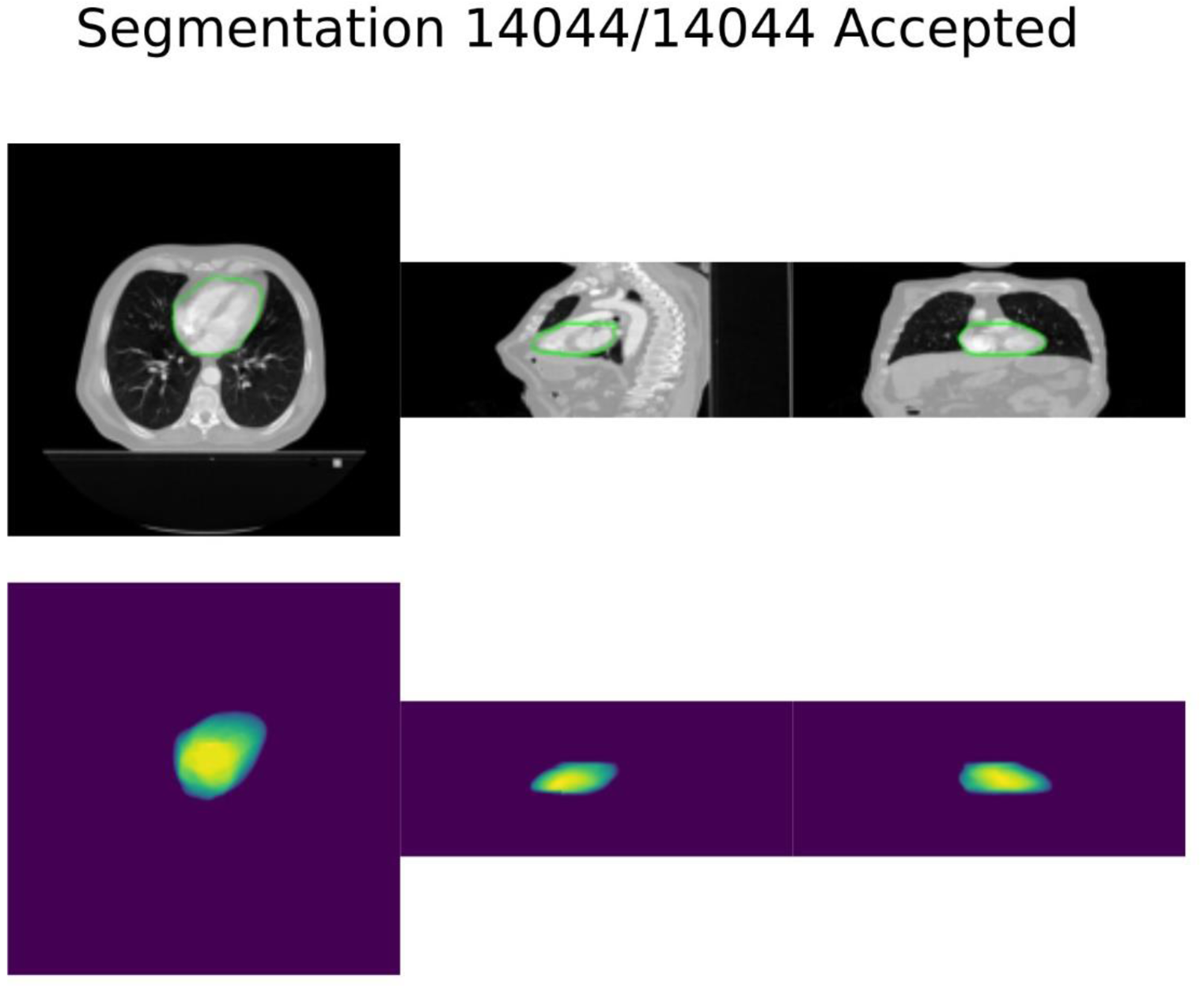
Example of software interface used for validation of the cardiac delineations. The top row displays the CT scan overlayed by the cardiac delineations in all three anatomical planes – axial, sagittal, and coronal (left to right) – taken at the center of mass of the delineation. The bottom row represents a rendering of the 3D cardiac delineation, where a more intense yellow color indicates a thicker section of heart. The user toggles through all patients and decides whether the delineation is acceptable, unacceptable, or undecided (which required further review with an oncologist).

**Figure A2.**
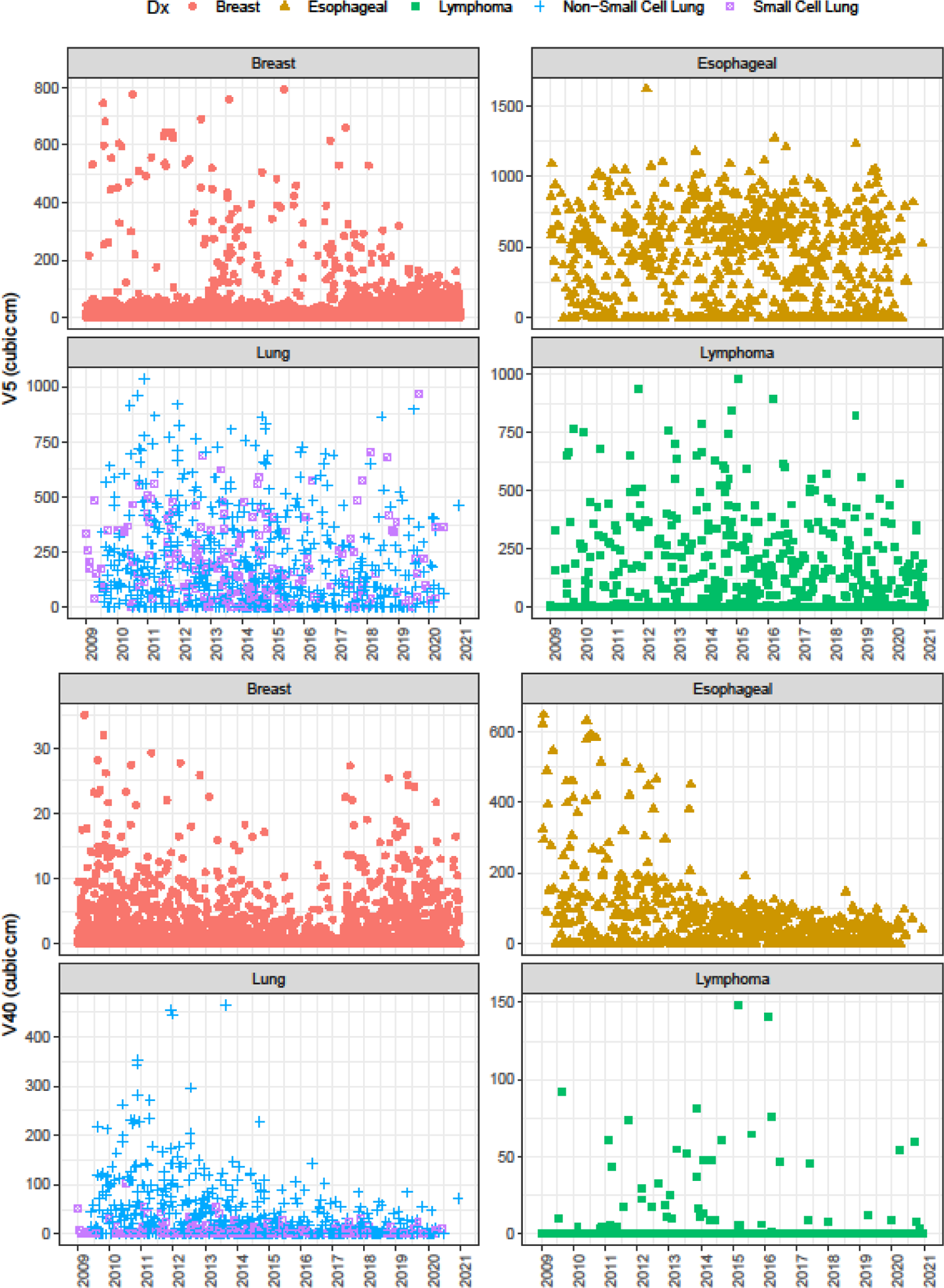
Scatter plots of heart volume receiving at least 5 Gy (V5) and heart volume receiving at least 40 Gy (V40) for each analysis group. Note that each panel has an independent y-axis scale, and in particular the breast cancer scale is relatively small.

